# Early Versus Delayed Add-on Therapy in Generalized Myasthenia Gravis: A Multicenter Real-World Cohort Study

**DOI:** 10.1101/2025.11.15.25340299

**Authors:** Menekse Oeztuerk, Niklas Huntemann, Lea Gerischer, Meret Herdick, Christopher Nelke, Frauke Stascheit, Sarah Hoffmann, Sophie Lehnerer, Maike Stein, Charlotte Schubert, Christiane Schneider-Gold, Steffen Pfeuffer, Heidrun H. Krämer, Franz Felix Konen, Thomas Skripuletz, Marc Pawlitzki, Christina B. Schroeter, Stefanie Glaubitz, Jana Zschüntzsch, Valerie Scherwietes, Andreas Totzeck, Tim Hagenacker, Andreas Meisel, Sven G. Meuth, Tobias Ruck

**Affiliations:** Ruhr University Bochum, BG University Hospital Bergmannsheil, Department of Neurology, Bochum, Germany; BG University Hospital Bergmannsheil, Heimer Institute for Muscle Research, Bochum, Germany; Department of Neurology, Medical Faculty, Heinrich Heine University Düsseldorf, Düsseldorf, Germany; Department of Neurology with Experimental Neurology, Charité –Universitätsmedizin Berlin, corporate member of Freie Universität Berlin, Humboldt-Universität zu Berlin, Berlin, Germany; Neuroscience Clinical Research Center, Charité – Universitätsmedizin Berlin, corporate member of Freie Universität Berlin, Humboldt-Universität zu Berlin, Berlin, Germany; Department of Neurology and Institute of Neuroimmunology and MS (INIMS), University Medical Center Hamburg-Eppendorf, Hamburg, Germany; Ruhr-University Bochum, Department of Neurology, St. Josef Hospital Bochum, Bochum, Germany; Department of Neurology, Justus Liebig University of Giessen, Giessen, Germany; Department of Neurology, Hannover Medical School, Hannover, Germany; Department of Neurology, University Medical Center Göttingen, Göttingen, Germany; Department of Neurology and Center for Translational Neuro– and Behavioral Sciences (C-TBNS), University Medicine Essen, Essen, Germany; Center for Stroke Research Berlin, Charité – Universitätsmedizin Berlin, corporate member of Freie Universität Berlin, Humboldt-Universität zu Berlin, Berlin, Germany

**Keywords:** myasthenia gravis, add-on therapy, treatment escalation, complement inhibition, FcRn inhibition

## Abstract

**Objective:** This study examined whether the timing of targeted add-on therapy initiation influences clinical outcomes in acetylcholine receptor (AChR) antibody-positive generalized myasthenia gravis (gMG), based on the hypothesis that earlier escalation may improve treatment response by intervening before structural or immunological consolidation occurs.

**Methods:** In this multicenter, retrospective real-world cohort study, 153 patients with AChR antibody-positive gMG were included from eight German tertiary centers. All received either complement C5 inhibitors (eculizumab, ravulizumab) or the FcRn antagonist efgartigimod as add-on therapy. Patients were grouped by treatment initiation within 24 months of diagnosis (Early Intensified Treatment; EIT) or later (Late Intensified Treatment; LIT). MG-ADL, QMG, and MG-QoL15 scores, as well as daily corticosteroid and pyridostigmine doses, were assessed at baseline and at 1, 3, and 6 months.

**Results:** The EIT group (n = 36) showed more pronounced and consistent clinical improvement. Significant differences emerged in maximum MG-ADL (p□=□0.013) and QMG (p□=□0.002) reductions. Patient-acceptable symptom states (MG-ADL ≤□2, QMG ≤□7) were more often reached with EIT (p□=□0.038, p□=□0.006). QMG worsening occurred only in the LIT group (n = 117) (p□=□0.021). Prednisone declined more steeply in EIT patients (p□=□0.001), alongside a trend toward reduced pyridostigmine use.

**Interpretation:** Initiating add-on therapy within two years of diagnosis was associated with stronger and more consistent clinical responses, fewer deteriorations, and a steeper reduction of treatment burden. These findings support timely escalation as a strategy to enhance both efficacy and tolerability in gMG care.

**Summary for Social Media If Published:** *What is the current knowledge on the topic?:* Complement and FcRn inhibitors have demonstrated efficacy in patients with AChR antibody-positive generalized myasthenia gravis (gMG) and are approved as add-on therapies in treatment-refractory disease. However, data guiding the optimal timing for their initiation in the treatment course remain limited.

*What question did this study address?:* This study investigated whether earlier initiation of add-on therapy improves clinical outcomes in AChR antibody-positive gMG. It compared patients escalated within 24 months of diagnosis to those treated later.

*What does this study add to our knowledge?:* Earlier treatment escalation was associated with significantly greater clinical improvements in MG-ADL and QMG scores, fewer symptom deteriorations, and steeper reductions in corticosteroid and pyridostigmine use, suggesting a potential benefit of timely intervention.

*How might this potentially impact the practice of neurology?:* The results support consideration of earlier escalation in the gMG treatment pathway. They may prompt re-evaluation of current stepwise approaches in favor of more proactive strategies.

*Suggested social media post:* Earlier add-on therapy in AChR+ gMG linked to better outcomes and lower treatment burden – real-world data suggest a benefit from timely escalation. Graphical Abstract:
Impact of Early Versus Late Initiation of Targeted Therapy in AChR-Positive Generalized Myasthenia Gravis. This graphical abstract illustrates the design and main findings of a multicenter, retrospective cohort study conducted across eight specialized MG centers in Germany. The study included 153 patients with acetylcholine receptor antibody-positive (AChR) generalized myasthenia gravis who received targeted add-on treatment with complement inhibitors (C5-I; eculizumab or ravulizumab) or an FcRn inhibitor (FcRn-I; efgartigimod). Participants were stratified based on the timing of escalation: those who initiated treatment within 24 months of diagnosis (Early Intensified Treatment; EIT) and those who escalated later (Late Intensified Treatment; LIT). Clinical outcomes were assessed using MG-ADL and QMG scores over a six-month period. Patients in the EIT group demonstrated more robust improvements in both functional and strength-based measures, with higher rates of clinically meaningful response and symptom resolution. The data support the notion that earlier introduction of targeted therapies may enhance treatment efficacy and improve patient outcomes in real-world settings. This figure was created with BioRender.com.

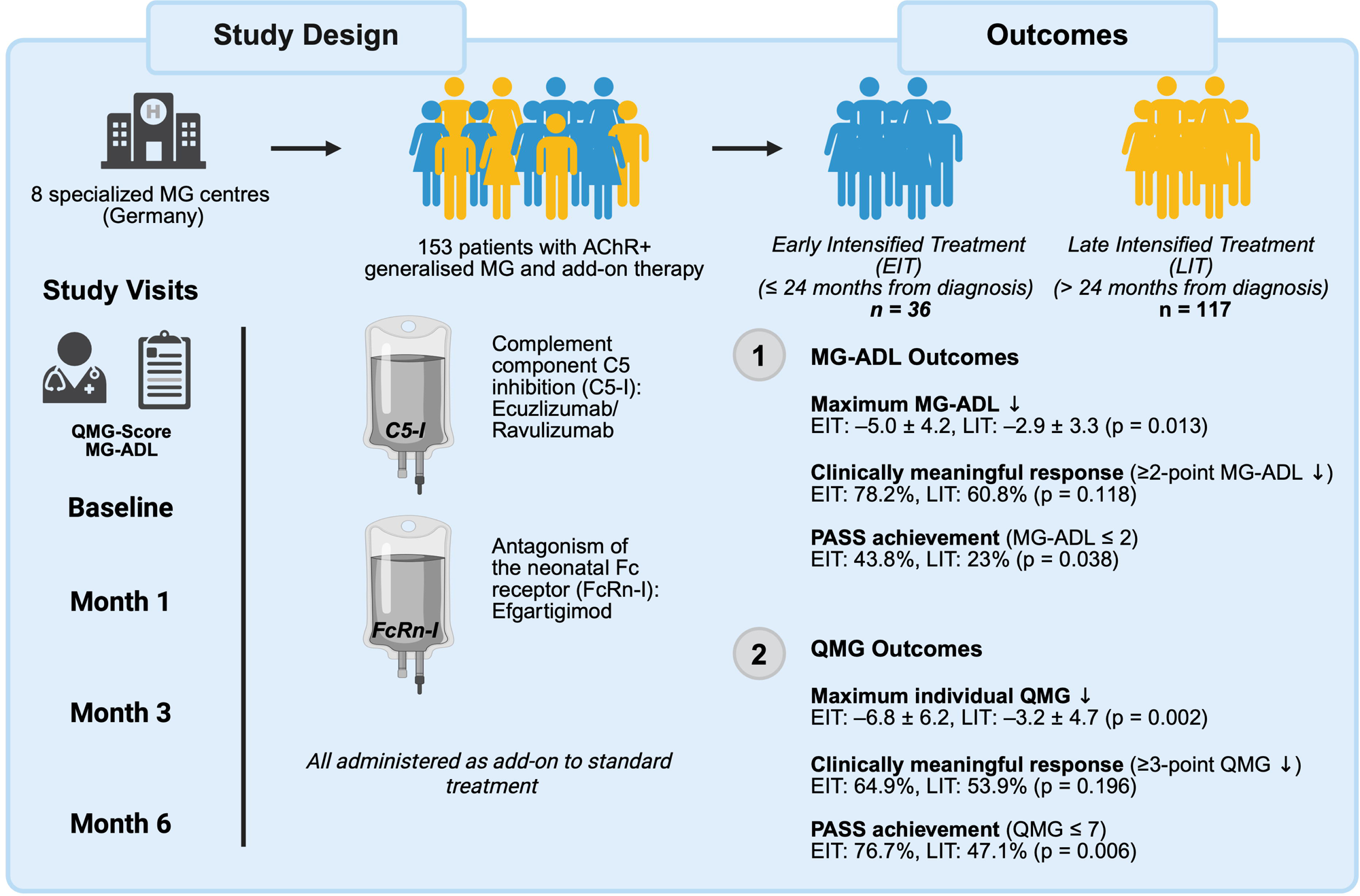

## Introduction

Myasthenia gravis (MG) is an antibody-mediated autoimmune disease that targets distinct components of the postsynaptic membrane at the neuromuscular junction (NMJ)^1^. Pathogenic autoantibodies, most commonly directed against the acetylcholine receptor (AChR), impede neuromuscular transmission and give rise to characteristic exertion-induced fluctuating weakness^2^. The clinical phenotype ranges from isolated ocular involvement to generalized forms with bulbar, limb and respiratory muscle weakness^3^. Although first-line immunosuppressive therapies achieve symptom control in a substantial proportion of patients, a clinically relevant subgroup experiences persistent disease activity, exacerbations, myasthenic crises or cumulative treatment toxicity^4–6^. These unmet clinical needs have driven the development of advanced immunotherapeutic strategies, among which two mechanistically distinct approaches have gained particular prominence. Experimental evidence has identified complement-mediated cytotoxicity as an effector mechanism in AChR antibody-positive MG, with deposition of the membrane attack complex at the NMJ driving structural endplate damage^7,8^. To counteract this mechanism, complement component C5 inhibition (C5-I) has been implemented through agents such as eculizumab^9^, ravulizumab^10^, and zilucoplan^11^, which interrupt terminal complement activation and thereby attenuate antibody-mediated structural injury at the NMJ. In parallel, antagonism of the neonatal Fc receptor (FcRn) has been pursued as a means of depleting pathogenic IgG. FcRn inhibitors (FcRn-I) like efgartigimod^12^, rozanolixizumab^13^ and nipocalimab^14^ disrupt IgG recycling, resulting in accelerated immunoglobulin clearance/degradation leading to significant reduction of serum IgG including pathogenic antibodies by about 70% and significant clinical improvement in respective phase III studies. Both classes are approved for use as add-on therapies in AChR antibody-positive generalized MG (gMG), following phase 3 trials in patients with inadequate response to standard immunosuppression.

Despite these advances, the optimal timing for initiating targeted add-on therapy remains clinically unstandardized. In both clinical trials and routine practice, escalation typically occurs after prolonged disease duration and failure of multiple immunosuppressants in so called “therapy-refractory” patients^15^.

However, sustained exposure to pathogenic autoantibodies has been associated with progressive disruption of the NMJ, including AChR loss, simplification of postsynaptic folds, and impaired synaptic transmission^16–18^. While it remains uncertain to which extent these structural changes can be reversed, delayed therapeutic escalation may limit clinical efficacy by missing a critical time window for intervention. In addition, prolonged immune activation may promote epitope spreading, potentially amplifying autoantibody diversity over time^19,20^. This consideration underscores the need to evaluate whether earlier initiation of add-on therapy, ideally before the onset of structural or immunological consolidation, might improve clinical outcomes.

In this multicenter, retrospective cohort study, we investigated the association between timing of treatment escalation and clinical outcomes in patients with AChR antibody-positive gMG. Patients were stratified according to whether targeted therapy was initiated within or beyond 24 months of diagnosis, a timeframe selected to reflect *real-world* treatment patterns, in which escalation typically follows an initial course of conventional immunosuppression^21,22^. We hypothesized that initiation within this period would be associated with a higher likelihood of clinical response and reduced symptom burden, reflecting a potential benefit of early therapeutic escalation.

## Material and Methods

### Study cohort

This retrospective, observational study included patients with gMG treated at eight German university hospitals between 2018 and 2024: University Hospital Bochum, Charité – Universitätsmedizin Berlin, University Hospital Düsseldorf, University Hospital Essen, University Hospital Gießen, University Medical Center Göttingen, University Hospital Hamburg-Eppendorf and Hannover Medical School. Diagnosis of MG was based on the presence of a typical clinical phenotype and the detection of autoantibodies directed against AChR, in accordance with national diagnostic standards^23^. All study sites applied unified procedures for diagnosis and treatment as outlined by the German Myasthenia Gravis Society. The dataset was sourced from the German Myasthenia Registry, and included demographic variables, antibody profiles, treatment regimens specific to MG, history of thymectomy and thymoma, comorbid conditions, adverse events (AE), as well as documented episodes of myasthenic crises and clinical exacerbation. Disease-related parameters were assessed and recorded by the treating neurologist at each site. All patients included in this study received standard therapy followed by escalation with an add-on treatment, either C5-I (eculizumab or ravulizumab) or FcRn-I (efgartigimod). Eligibility required an age of 18 years or older at the time of treatment initiation. Standard therapy was defined as treatment with corticosteroids, pyridostigmine, thymectomy when indicated and conventional immunosuppressive agents such as azathioprine, mycophenolate mofetil, methotrexate, cyclosporine A, or tacrolimus. The cohort included in this study has previously been analyzed and published in the context of a different objective^24^. The study followed the STROBE reporting guideline^25^.

### Definitions and Outcome Measures

Patients were grouped according to the interval between diagnosis and initiation of targeted add-on therapy. Treatment escalation within 24 months of diagnosis was defined as Early Intensified Treatment (EIT); escalation beyond this threshold was classified as Late Intensified Treatment (LIT). MG Activities of Daily Living (ADL), quantitative MG (QMG) scores and MG-specific quality of life (MG-QoL15) were assessed at baseline (BL) and at follow-up visits 1 (14-45 days post-BL), 3 (60 – 120 days post-BL) and 6 months after BL (150 – 210 days post-BL)^26–28^.

Endpoints included both the maximum and a clinically meaningful improvement in MG-ADL and QMG, point-specific changes in both MG-ADL and QMG at months 1, 3, and 6, and the proportions of patients reaching predefined clinical thresholds. Clinically meaningful improvement was defined as a reduction of ≥2 points in MG-ADL or ≥3 points in QMG. The Patient Acceptable Symptom State (PASS) was defined as an MG-ADL score ≤2, QMG score ≤7 or a MGl7lQoL15 ≤ 8, in accordance with established thresholds^29^. Minimal symptom expression (MSE) was defined as an MG-ADL score ≤1^30^. Symptom worsening was defined as any increase in MG-ADL or QMG scores relative to BL. All definitions and thresholds were prespecified before data analysis.

### Standard protocol approvals, registrations, and patient consents

All patients provided informed consent within the framework of the German Myasthenia Registry (https://dmg.online/myasthenie-register). The study was conducted in accordance with the Declaration of Helsinki and followed the STROBE reporting guidelines. It was prospectively registered in the WHO-accredited German Clinical Trials Register (DRKS00024099). Ethical approval was granted by the ethics committee of Charité – Universitätsmedizin Berlin (EA1/214/18); for the Hannover site, patient inclusion was covered by a separate ethics vote (9741_BO_S_2021). All data were pseudonymized and collected retrospectively.

### Statistical analysis

Statistical Analysis was performed using Prism10 (GraphPad Software, Inc.). Continuous data are presented as mean (± standard deviation (SD)). Differences between groups were analysed using a two-tailed, unpaired t-test for quantitative variables and two-tailed Fisher’s exact test for categorical variables. For comparison of the best individual response with BL scores, a paired t-test was applied. Differences were considered statistically significant with the following P-values: * *p* < 0.05, ** *p* < 0.01, *** *p* < 0.001.

## Results

A total of 153 patients with gMG who were treated with either C5-I (eculizumab or ravulizumab) or FcRn-I (efgartigimod) were included in the analysis. Of these, 36 patients initiated treatment within 24 months of diagnosis and were assigned to the EIT group.

The remaining 117 patients received escalation therapy beyond 24 months from diagnosis and were assigned to the LIT group. BL demographical and clinical characteristics for both groups are presented in Table 1. The proportion of female patients was similar between cohorts, with 66.7% (24 patients) in the EIT group and 70.1% (82 patients) among those with delayed treatment. Mean age at treatment initiation was 49.1□±□21.4 years among EIT individuals and 54.7□±□19.2 years in those treated later. Age at diagnosis was similar, averaging 48.1□±□21.2 and 46.0□±□20.8 years, respectively. The distribution of early-onset MG (defined as onset before age 50) was also well balanced, occurring in 55.6% (20 patients) of the earlier-treated individuals and 53.8% (63 patients) of those escalated at a later stage. Disease severity at BL was similar across groups: Mean MG-ADL scores were 9.0□±□5.0 in the early group and 8.8□±□4.4 in the late group. Corresponding QMG scores averaged 11.3□±□5.6 and 12.5□±□6.8, respectively. MG-QoL15 scores at BL were 27.0□±□12.8 in the early group and 29.2□±□13.4 in the late group.

**Table 1:** Baseline characteristics for patients undergoing early or late therapy intensification (total n = 153). Table 1 presents BL characteristics for patients with early or late add-on treatment initiation. Patients were stratified based on the interval between diagnosis and treatment escalation, with early initiation defined as therapy started within 24 months of diagnosis. Baseline (BL) was set as the date of first administration of a C5 inhibitor or FcRn antagonist. Disease duration refers to the time from symptom onset to BL. Early-onset MG (EOMG) was defined as symptom onset before the age of 50. Comparisons between groups were performed using two-sided Student’s t-tests for quantitative variables and Fisher’s exact tests for qualitative variables. A p-value <□0.05 was considered statistically significant. *Ab: antibody; AChR: acetylcholine receptor; BL: baseline; d: days; EOMG: early onset myasthenia gravis; FcRn: neonatal Fc receptor; ISTs: immunosuppressive therapies; max: maximum; MG: myasthenia gravis; MG-ADL: Myasthenia Gravis Activities of Daily Living; MGFA: Myasthenia Gravis Foundation of America; MG-QoL15: Myasthenia Gravis Quality of Life 15-Item Questionnaire; min: minimum; MuSK: muscle-specific tyrosine kinase; n: number; QMG: Quantitative Myasthenia Gravis; SD: standard deviation, w: weeks; y: years*.

| Characteristics | Early | Late | p-value |
| --- | --- | --- | --- |
| Patients, n | 36 | 117 |  |
| Female patients, n (%) | 24 (66.7) | 82 (70.1) | 0.685 |
| Age at baseline, y, mean (SD) | 49.1 (21.4) | 54.7 (19.2) | 0.168 |
| Age at diagnosis, y, mean (SD) | 48.1 (21.2) | 46 (20.8) | 0.643 |
| EOMG, n (%) | 20 (55.6) | 63 (53.8) | > 0.999 |
| **Disease duration, y, mean (SD)** | **1.3 (1)** | **9.6 (8.4)** | **< 0.001** |
| History of thymectomy, n (%) | 20 (55.6) | 74 (63.2) | 0.438 |
| Confirmed thymoma, n (%) | 8 (22.2) | 17 (14.3) | 0.305 |
| **Total number of previous ISTs, mean (min-max)** | **1.7 (1-3)** | **2.9 (1-7)** | **< 0.001** |
| **Prednisolone dose at BL, mg/d, mean (SD)** | **24.7 (23.6)** | **8.8 (11.2)** | **< 0.001** |
| **Pyridostigmine dose at BL, mg/d, mean (SD)** | 397 (194) | 364.1 (176) | 0.675 |
| QMG score at BL, mean (SD) | 11.3 (5.6) | 12.5 (6.8) | 0.291 |
| MG-ADL at BL, mean (SD) | 9 (5) | 8.8 (4.4) | 0.746 |
| MG-QoL15 at BL, mean (SD) | 26.6 (13.2) | 28.3 (13.6) | 0.419 |
| Antibody status |  |  |  |
| seronegative, n (%) | 3 (8.3) | 1 (0.9) | 0.041 |
| seropositive, n (%) | 33 (91.7) | 116 (99.1) | 0.041 |
| anti-AChR-Ab, n (%) | 33 (91.7) | 116 (99.1) | 0.041 |
| anti-LRP4-Ab, n (%) | 2 (5.6) | 2 (1.7) | 0.236 |
| anti-Titin-Ab, n (%) | 5 (13.9) | 19 (16.2) | > 0.999 |
| anti-MuSK-Ab, n (%) | 0 (0) | 1 (0.9) | > 0.999 |
| MGFA at BL, n (%) |  |  |  |
| I | 0 (0) | 0 (0) | > 0.999 |
| IIA | 3 (8.3) | 10 (8.5) | > 0.999 |
| IIB | 4 (11.1) | 16 (13.7) | 0.785 |
| IIIA | 9 (25) | 40 (34.2) | 0.414 |
| IIIB | 13 (36.1) | 34 (29.1) | 0.417 |
| IVA | 2 (5.6) | 6 (5.1) | > 0.999 |
| IVB | 5 (13.9) | 10 (7.5) | > 0.347 |
| V | 0 (0) | 1 (0.9) | > 0.999 |

### MG-ADL outcomes

Following initiation of therapy with either C5-I or FcRn-I, those in the EIT cohort showed a marked reduction in MG-ADL scores as early as one month post-treatment, with a mean change of –3.0□±□4.1 points compared to BL (p = 0.0005). This improvement was sustained over time, with mean reductions of –3.4□±□3.6 (p = 0.0004) and –4.2 ± 3.7 points (p < 0.0001) observed at months 3 and 6, respectively. In contrast, the LIT group exhibited a more gradual and less pronounced reduction in MG-ADL, with corresponding changes of –1.5□±□3.6 at month 3 (p = 0.001) and – 1.5□±□4.7 at month 6 (p = 0.0012). The maximum individual improvement in MG-ADL during the entire observation period was –5.0□±□4.2 points among the EIT cohort and –2.9□±□3.3 points in the LIT group (p□=□0.013), reflecting a more robust treatment effect when escalation was introduced earlier. PASS was reached by 43.8% of EIT patients, while this threshold was achieved by 23% of LIT patients (p□=□0.038). Similarly, the proportion of patients achieving a clinically meaningful MG-ADL improvement tended to be higher in the EIT group (78.2%) compared to those with delayed escalation (60.8%) (p□= 0.118). MSE was documented in 21.9% of EIT and 14.9% of LIT patients (p□=□0.41). Based on the highest individual MG-ADL response, clinical deterioration was reported in 3.2% of the early and 6.8% of the late treatment groups (p = 0.665) (Fig. 1A).

**Figure 1.**
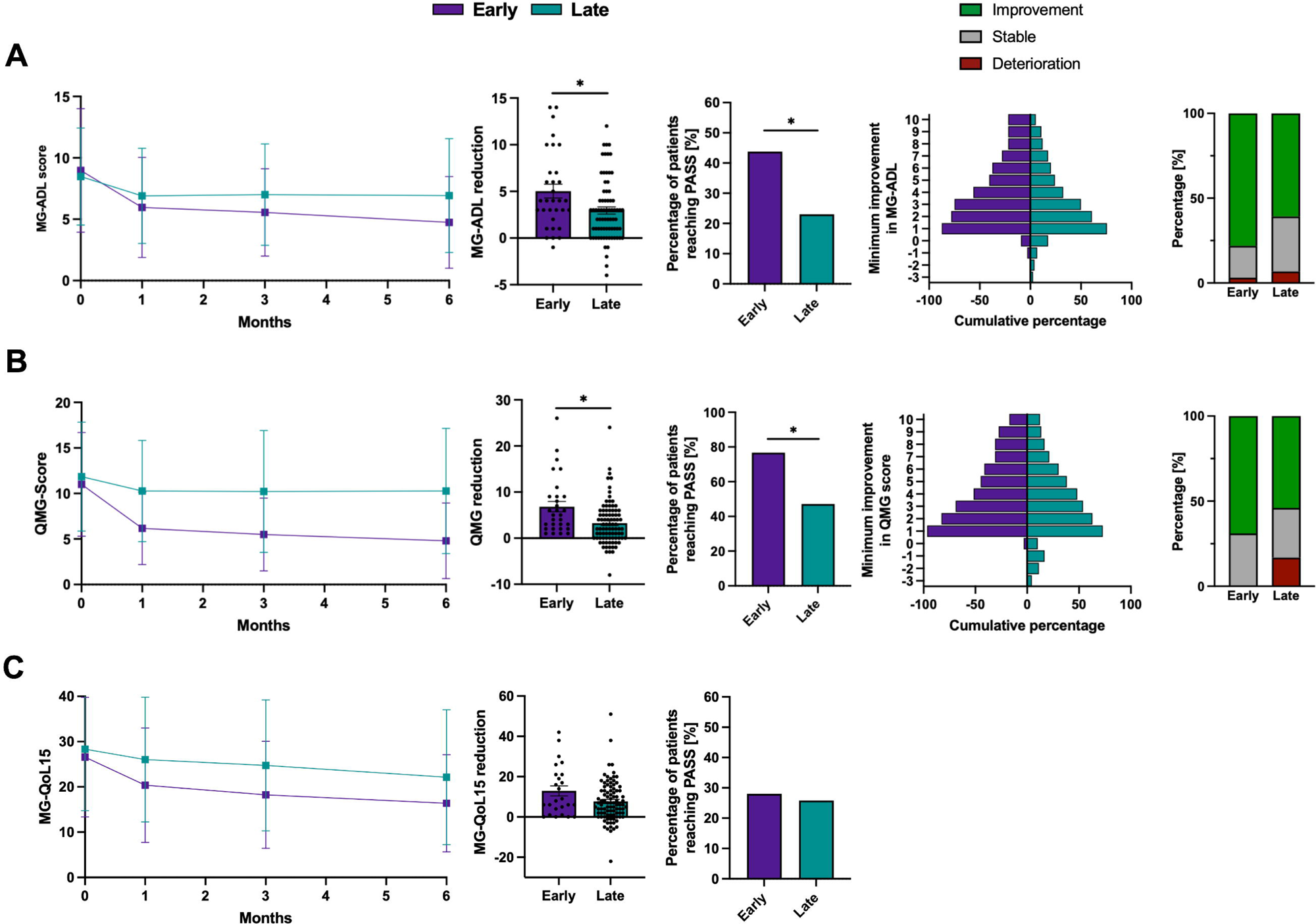
Clinical outcome parameters in patients undergoing early (n = 36) versus late (n = 117) treatment escalation. (**A**) The left panel displays mean MG-ADL scores at BL and at months 1, 3, and 6 following treatment initiation, stratified by early (purple) and late (green) escalation groups. The individual maximal reduction in MG-ADL scores is presented in the centre-left, while the proportion of patients achieving a PASS based on the MG-ADL score as defined by Mendoza et al. (MG-ADL ≤ 2 is depicted in the centre. The centre-right diagram shows the minimal improvements in MG-ADL score. The right diagram depicts the proportion who experienced a MG-ADL deterioration (red), a stable MG-ADL (reduction of 0-1 points, grey), or a relevant improvement (MG-ADL reduction ≥ 2 points; green). Figures (**B**) illustrate the average QMG scores at months 1, 3, and 6 post-BL (left), the maximum decrease in QMG score (centre-left), the percentage of patients meeting PASS criteria based on the QMG score (QMG ≤ 7; centre), and the minimum improvement in total QMG scores (centre-right). The right graph shows the proportion of patients with a QMG deterioration (red), a stable QMG (QMG reduction of 0-2 points, grey), or a significant improvement (QMG reduction ≥ 3 points; green). In **(C)**, mean MG-QoL15 scores (left), the individual best MG-QoL15 response as well as the percentage of patients meeting PASS criteria based on the QMG score (QMG ≤ 8; right) are depicted. Error bars represent the mean with SD. Quantitative variables were analysed using a two-sided Student’s t-test A p-value < 0.05 was considered statistically significant. *BL; baseline; d: day; MG-ADL: Myasthenia Gravis Activities of Daily Living; MG-QoL15: Myasthenia Gravis Quality of Life 15-Item Questionnaire; PASS: Patient Acceptable Symptom State; QMG: Quantitative Myasthenia Gravis; SD: Standard deviation*.

### QMG outcomes

Similar trends were observed when evaluating the QMG score. One month after initiation of therapy, the EIT group exhibited a mean QMG reduction of –4.8 ± 4.8 points (p = 0.001), while the LIT cohort showed a change of –1.7 ± 6 points (p < 0.001). Continued improvement was observed in EIT individuals, with reductions of – 5.5 ± 4.9 points at month 3 (p = 0.001) and –6.2 ± 5.1 points at month 6 (p < 0.001), indicating a progressive benefit over time. In contrast, the LIT group showed a flatter response profile, with reductions of –1.7 ± 7.2 (p = 0.009) and –1.6 ± 7.4 points (p = 0.019) at months 3 and 6, respectively. The maximum individual QMG reduction during follow-up was –6.8□± 6.2 in LIT individuals and –3.2 ± 4.7 in the EIT group (p = 0.002).

Based on the best individual QMG score, 76.7% of EIT patients and 47.1% of patients of the LIT cohort achieved a PASS (p = 0.006) (Fig. 1B). A clinically meaningful QMG response was observed in 64.9% of EIT patients and 53.9% of those treated later (p = 0.196) (Fig. 1B). Symptom worsening, defined as any deterioration in QMG compared to BL, was recorded in 0% of EIT individuals and 16.9% of the LIT group (p = 0.021) (Fig. 1B).

### MG-QoL15 outcomes

One month after the first C5-I/ FcRn-I dose, EIT patients showed a mean MGl7lQoL15 reduction of –6.2 points (from 26.6 ± 13.2 to 20.4 ± 12.6; p = 0.0363), whereas the LIT cohort improved by –2.3 points (28.3 ± 13.6 to 26.0 ± 13.8; p = 0.0006). At months 3 and 6, the EIT group achieved mean decreases of –8.4 (p = 0.06) and –10.2 points (p = 0.0008), respectively, compared with –3.6 (p = 0.0004) and –6.2 points in the LIT group (p = 0.0036). The greatest individual improvement recorded during followl7lup was –12.9 ± 12.3 points in the EIT cohort versus –7.7 ± 10.0 points after delayed initiation (p = 0.079). A PASS was attained by 28.0% of patients in the EIT group and by 25.8% of those treated later (p = 0.803) (Fig. 1C).

### Concomitant medication

In the EIT cohort, the mean daily prednisone dose fell by 14.9% after one month, 27% at month 3 and 40.5% at 6 months after treatment initiation (Fig. 2A). In contrast, individuals in the LIT cohort exhibited only modest reductions, with decreases of 4.1%, 12.8%, and 18.8% at months 1, 3, and 6, respectively. Patients escalated early exhibited a markedly greater maximal reduction in prednisolone compared with the LIT cohort (EIT: –13.5 ± 20 mg; LIT: –3.5 ± 8 mg; p = 0.001).

**Figure 2.**
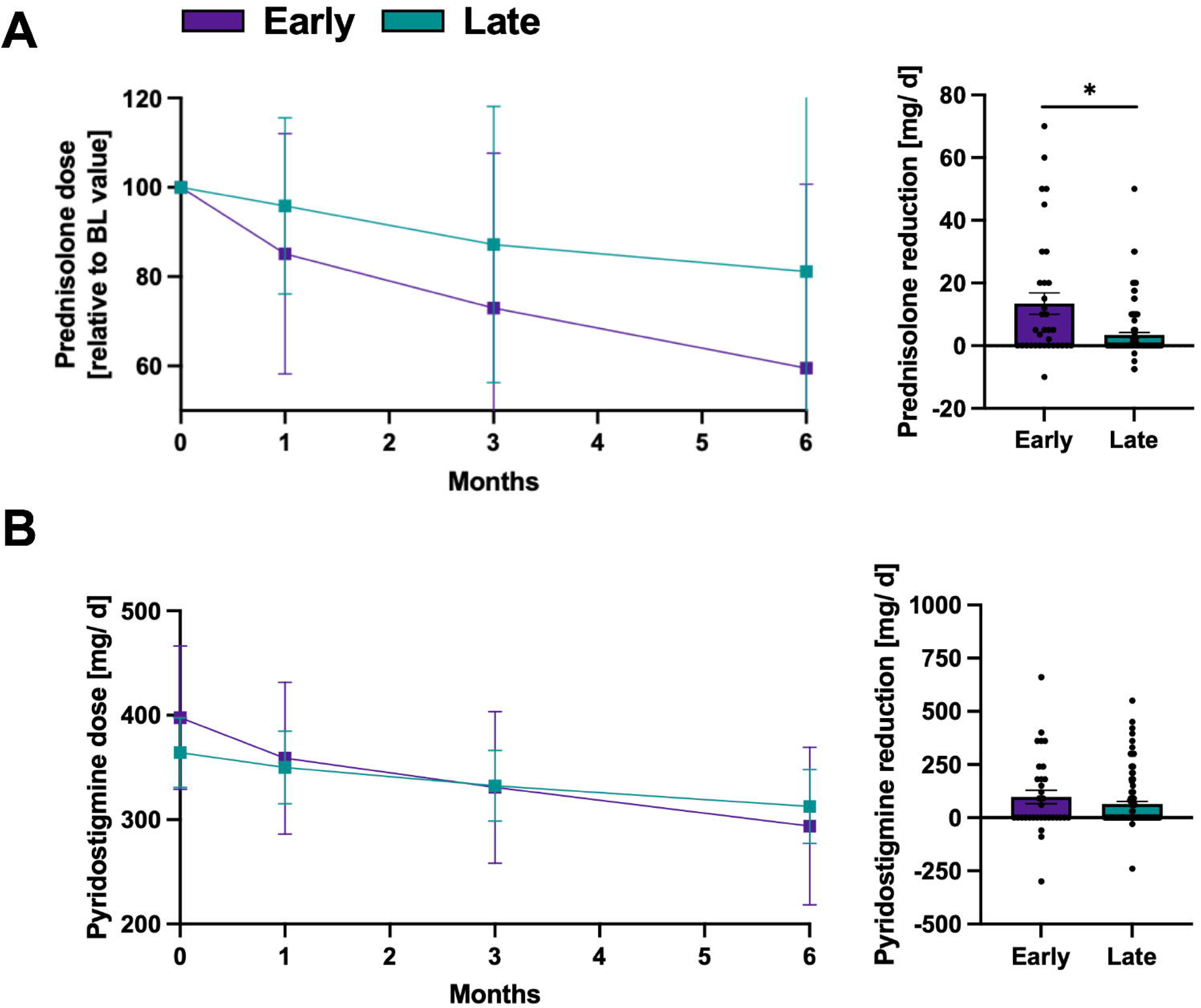
Changes of the concomitant medication in patients undergoing early (n = 36) versus late (n = 117) treatment escalation. (A) Mean prednisolone doses at BL and at months 1, 3, and 6 following initiation of add-on therapy in patients treated early (purple) or late (green) after diagnosis (left). The right panel shows the maximal reductions of daily prednisolone doses within the first six months after treatment escalation. (B) Average pyridostigmine doses after treatment intensification (left). Maximal reduction of pyridostigmine for patients with early and late treatment escalation. Error bars represent the mean with SD. Quantitative variables were analysed using a two-sided Student’s t-test A p-value < 0.05 was considered statistically significant. *BL: baseline; SD: Standard deviation*.

Pyridostigmine usage followed the same trend. EIT patients reduced their average daily intake from 397 ± 194 mg to 358.8 ± 205 mg (month 1), 331.1 ± 205 mg (month 3), and 293.9 ± 213 mg (month 6), achieving a maximum individual reduction of –97.6 ± 180.4 mg (Figure 2B). The LIT cohort declined from 364.1 ± 176 mg to 349.9 ± 183 mg, 332.5 ± 178 mg and 312.6 ± 187 mg, respectively, with a maximal change of –64.8 ± 123.7 mg (p = 0.381 compared to the EIT cohort) (Fig. 2B).

In further analyses, we applied a 12-month cut-off, reasoning that an even earlier start of therapy might yield more pronounced differences. The results followed the same overall tendency, however, the small patient cohort of EIT patients constrains the robustness of these findings (Supplemental Figure 1). Examining the effect of EIT on therapeutic response separately for the two drug classes, C5-I and FcRn-I, revealed that an earlier initiation of therapy tended to be associated with better clinical outcomes in both groups (Supplemental Figures 2 and 3).

## Discussion

Although targeted therapies have expanded the treatment landscape of gMG, it remains unclear whether the timing of escalation influences clinical outcomes. While regulatory approvals and clinical trials have established efficacy in treatment-refractory disease, they provide no guidance on when these agents should ideally be introduced. To the best of our knowledge, no *real-world* data have systematically addressed whether earlier initiation of add-on therapy leads to improved response. By analyzing a large multicenter cohort, this study provides data to support clinical decision-making and optimize the timing of therapeutic escalation.

Patients were stratified by a 24-month threshold from diagnosis to initiation of targeted therapy. This cut-off reflects pre-2023 treatment paradigms, when escalating to advanced therapies was typically deferred until a treatment-resistant phenotype emerged, as codified in earlier German neurological guidelines^23^. Prior to the 2023 guideline revision, which introduced disease activity assessment (e.g. MG-ADL, QMG scores) as a trigger for early intensification, clinical practice favored a stepped-care approach^31^. In most cases, escalation does not occur at diagnosis. Patients are typically started on standard immunosuppressive agents, such as azathioprine or mycophenolate mofetil, which require several months before clinical benefit becomes apparent^32^. As a result, treatment intensification often takes place only after a prolonged course of suboptimal disease control^33^. Additional delays may arise from hesitancy toward newer treatment options or access-related barriers.

In this *real-world* cohort, earlier initiation of targeted therapy was associated with more pronounced and sustained improvements across multiple clinical measures. Notably, this effect emerged despite the absence of major differences in BL demographic or disease characteristics between groups. The EIT group showed earlier and significantly greater improvements in MG-ADL and QMG scores compared to those treated later. While both groups experienced clinical benefit over time, responses in the EIT group appeared more consistent and more frequently met established thresholds for meaningful improvement and symptom resolution. MG-ADL was selected as the primary outcome measure given its established role in clinical trials and regulatory approvals for targeted therapies in gMG^9–13^. As a patient-reported outcome measure, MG-ADL quantifies the functional impact of generalized muscle weakness on everyday activities and serves as a clinically relevant indicator of patient-perceived disease burden. Additional outcome measures, including QMG-based response thresholds, PASS, and MSE, were selected to complement MG-ADL by offering objective, clinician-assessed indices of neuromuscular strength and disease control. Statistically significant differences were observed for MG-ADL-based PASS (p = 0.038), QMG-based PASS (p = 0.006), maximal MG-ADL improvement (p = 0.013), and symptom worsening in QMG, which occurred exclusively in the LIT group (p = 0.021). While not all additional differences reached statistical significance, patients in the EIT group more frequently achieved key clinical benchmarks, including clinically meaningful MG-ADL and QMG improvement, and MG-ADL-defined MSE. Moreover, MG-ADL deterioration was observed less frequently in the EIT group, reinforcing the trend toward more stable disease control under earlier escalation. Although the maximum individual QMG reduction was numerically greater in the LIT group, this appears to reflect variability in individual treatment responses rather than a systematic effect of later initiation. Given that overall mean improvements and response rates favored EIT and BL disease severity was similar, this isolated finding does not alter the general trend observed across the cohort.

A more pronounced reduction in concomitant medication burden was observed among patients of the EIT group. This subset achieved significantly greater reductions in daily prednisolone dosage over the 6-month observation period. Importantly, higher BL doses in the EIT group reflect typical clinical practice, where corticosteroids are often initiated at higher levels in the early disease course. While this naturally allows for a larger numerical reduction, the consistent tapering trend nonetheless suggests that EIT may support more effective steroid withdrawal under targeted therapy. In addition to the absolute decrease, the data also indicate a relatively steeper decline in daily corticosteroid use over time in the EIT group, indicating that early intervention may enable more efficient control of disease progression. A similar trend was observed for pyridostigmine, pointing to a broader reduction in symptomatic treatment requirements.

Our findings are consistent with the still limited body of published evidence to date. For instance, a post-hoc analysis of the CHAMPION-MG trial demonstrated that initiating eculizumab ≤ 2 years after diagnosis led to significantly greater improvements in MG-ADL scores than initiation at later disease stages^34,35^. Similarly, Nagane et al. performed a retrospective comparison of standard oral prednisolone with an early aggressive regimen combining plasmapheresis and high dose intravenous methylprednisolone^36^. This analysis displayed that EIT led to superior clinical outcomes and permits lower maintenance prednisolone doses^36^. Evidence from the Japanese MG registry reinforced these observations since early fast-acting treatment was associated with a higher probability of achieving minimal-manifestation status on ≤ 5 mg/day prednisolone^37^. A Swedish retrospective cohort study showed that B-cell-depleting therapy with rituximab, when started at disease onset rather than in refractory gMG, was linked to a more rapid remission, reduced need for rescue therapy, and lower concomitant medication burden. The authors therefore advocated for early use of rituximab in the disease course^38^.

Building on the evidence from histopathological studies, one plausible explanation for the observed advantage of early initiation lies in the structural vulnerability of the NMJ in MG. Prolonged exposure to pathogenic antibodies has been shown to trigger complement activation, resulting in deposition of the membrane attack complex at the NMJ and subsequent structural degradation, including loss of acetylcholine receptors and simplification of postsynaptic folds^7,8,39^. These changes may be only partially reversible, potentially limiting the efficacy of therapeutic interventions when introduced at a later disease stage. Early therapeutic escalation, particularly with agents that inhibit complement activity or reduce IgG levels, may help preserve NMJ integrity before irreversible damage has occurred^18,40^. Beyond structural considerations at the NMJ, immunological mechanisms may also contribute to the clinical benefit associated with earlier therapeutic escalation. One such mechanism is epitope spreading, wherein the autoimmune response broadens over time to include additional antigenic targets beyond the initial epitope^41,42^. This process, driven by ongoing immune activation and tissue injury, has been described in various autoimmune conditions and is thought to play a role in disease progression and treatment resistance^19,43^. In MG, it has been particularly observed in MuSK-positive cases, where antibody reactivity may evolve to encompass multiple regions of the same autoantigen^20,44^. Although this process is less well characterized in AChR-positive MG, findings from experimental models and patient sera suggest that antigenic targets may diversify over time, potentially amplifying the autoimmune response^45^. In this light, the more consistent treatment effects observed in patients with earlier escalation may not only reflect preserved endplate integrity, but also a more contained and modifiable autoimmune process. Our clinical findings of stronger and more consistent responses among patients treated earlier are consistent with this model and lend support to the hypothesis that there is a window of opportunity in which targeted therapies are most effective.

### Limitations

Several methodological constraints should be considered when interpreting the results. As a retrospective analysis, it is subject to the usual constraints of non-randomized observational data, including potential confounders that cannot be fully controlled. All patients included in this study were managed at tertiary referral centers. While this may limit transferability to general neurology settings, it reflects current practice conditions: the use of C5-I and FcRn-I is largely concentrated in specialized centers. The study population thus corresponds to the group in which these treatments are most commonly initiated. Notably, BL characteristics including age, disease duration, and prior MG-specific treatment aligned with those observed in another large German MG study^46^. A further limitation concerns the timing-related resolution of clinical assessments. Scores were documented at fixed intervals, namely at 1, 3 and 6 months after treatment initiation, which may not have captured each patient’s optimal treatment response, particularly in the context of symptom fluctuation or delayed onset of action. To address this, we analyzed each patient’s best individual response within the observation period, offering a pragmatic estimate of peak clinical effect. Still, more detailed monitoring might have offered deeper insight into individual treatment trajectories.

An important limitation concerns corticosteroid exposure. Patients in the EIT group started on higher BL prednisolone doses, which reflects common practice early in the disease course. Higher corticosteroid exposure at BL could have contributed to the initial improvement in MG-specific outcome measures, independent of add-on therapy effects. However, given that outcome differences between groups persisted across multiple parameters and timepoints, a steroid-driven explanation alone appears insufficient. Nevertheless, the influence of BL corticosteroid intensity cannot be entirely disentangled and represents a relevant limitation.

## Conclusions

This *real-world*, multicenter cohort study examined whether the timing of add-on therapy influences clinical outcomes in AChR antibody-positive gMG. The analysis suggests that initiating treatment within 24 months of diagnosis may be associated with greater and more sustained clinical benefit than delayed escalation. These results support the concept of a time-sensitive therapeutic window during which targeted intervention may be most effective. Early initiation was also associated with a more favorable reduction in concomitant medication, pointing to a potential for improved tolerability alongside clinical benefit.

Prospective trials are needed to clarify the underlying mechanisms and validate these observations. This may include integration of serological and histopathological markers to identify patients most likely to benefit from early escalation. In addition, digital monitoring approaches such as wearable sensors or remote symptom tracking could offer continuous assessment beyond scheduled visits and provide a more holistic view of individual treatment dynamics.

## Acknowledgements

We thank the patients and their families for their valuable contribution and support. This work was funded by the Else Kröner-Fresenius-Stiftung to CN (2023_EKEA.38). Further, this work was supported by the internal research funding program of the Heinrich Heine University to MO and CN. Further study funding by the DGM (Deutsche Gesellschaft für Muskelkranke) to TR and to CN, by the German Research Foundation (DFG, to CN: NE 2774/2-1, to TR: 549557400/RU 2169/5-1, 417677437/GRK2578).

## Author contributions

MO, NH, AM, and TR designed the study and methods. Formal analysis was done by NH and TR. Clinical data were provided by MO, NH, LG, MH, CN, FS, SH, CBS, SL, MS, CS, CS-G, SP, HHK, FFK, TS, MP, SG, JZ, VS, AT, TH, SGM, AM, and TR. Resources were provided by AM and TR. MO, NH and TR wrote the original draft. LG, MH, CN, FS, SH, CBS, SL, MS, CS, CS-G, SP, HHK, FFK, TS, MP, SG, JZ, VS, AT, TH, SGM, AM reviewed and edited the manuscript. Figures were created by MO, NH and TR. Supervision by AM and TR. Guarantors of the study are AM and TR.

## Potential Conflicts of Interest

The authors report no conflicts of interest.

## Data availability

All analyzed data are presented in the manuscript and available on reasonable request from qualified investigators.

## Figure legends

**Supplemental Figure 1.** Outcome parameters in patients undergoing treatment escalation within 13 months of diagnosis versus thereafter. This figure depicts the trajectory during the first 6 months after therapy escalation as well as the individual best response for **(A)** the MG-ADL, **(B)** QMG score, **(C)** daily prednisolone and **(D)** daily pyridostigmine dose. Error bars represent the mean with SD. Quantitative variables were analysed using a two-sided Student’s t-test A p-value < 0.05 was considered statistically significant. *BL, baseline; MG-ADL, Myasthenia Gravis Activities of Daily Living; QMG, Quantitative Myasthenia Gravis; SD, standard deviation*.

**Supplemental Figure 2.** Clinical outcomes after early (within 24 months of diagnosis) versus late (thereafter) initiation of complement C5 inhibition. The figure shows the course of key endpoints in patients treated with a C5 inhibitor, contrasting early (purple) and late (green) escalation. **(A)** depicts MG-ADL outcomes, showing mean scores at BL and at months 1, 3 and 6 (left), the maximal individual reduction in MG-ADL (centre-left), the proportion of patients reaching a PASS (MG-ADL score ≤ 2; centre), and the proportion achieving a clinically meaningful improvement of at least two points (centre-right). The right diagram depicts the proportion who experienced a MG-ADL deterioration (red), a stable MG-ADL (reduction of 0-1 points, grey), or a relevant improvement (MG-ADL reduction ≥ 2 points; green). **(B)** provides the corresponding QMG analyses, presented as mean scores within the first six months after treatment initiation, the individual maximal reduction and the proportion meeting PASS criteria (QMG ≤ 7 points). The right graph shows the proportion of patients with a QMG deterioration (red), a stable QMG (QMG reduction of 0-2 points, grey), or a significant improvement (QMG reduction ≥ 3 points; green). **(C)** illustrates the MG-QoL15, displaying mean scores over time (left) together with the best individual improvement and the proportion attaining PASS (score ≤ 8; right). In **D**, prednisolone dosing is depicted, showing mean daily prednisone doses at BL and at months 1,3, as well as 6 (left), and the greatest individual dose reduction within six months (right). **(E)** presents analogous data for pyridostigmine, with mean daily doses over time (left) and the maximal individual reduction achieved (right). Error bars represent mean ± SD. Groups were compared with two-sided Student’s t-tests; p < 0.05 was considered significant. *BL, baseline; MG-ADL, Myasthenia Gravis Activities of Daily Living; QMG, Quantitative Myasthenia Gravis; PASS, Patient-Acceptable Symptom State; SD, standard deviation*.

**Supplemental Figure 3.** Clinical outcomes following early (within 24 months of diagnosis) versus late (thereafter) initiation of the FcRn antagonist efgartigimod. This figure displays key clinical and pharmacological parameters in patients escalated to efgartigimod, comparing early (purple) and late (green) initiation. Panel **A** depicts MG-ADL dynamics, presenting mean scores at BL and at months 1, 3 and 6 (left), each patient’s maximal MG-ADL reduction (centre-left), the proportion achieving PASS (score ≤ 2; centre) and patients attaining a ≥ 2-point improvement (centre-right). The right graph shows the proportion of patients with a MG-ADL deterioration (red), a stable MG-ADL (MG-ADL reduction of 0-1 points, grey), or a significant improvement (MG-ADL reduction ≥ 2 points; green). **(B)** depicts the analogous QMG analyses: mean scores over the first six months, maximal individual reductions, and the proportion fulfilling PASS criteria (QMG ≤ 7). The right diagram shows the distribution of patients with QMG worsening (red), stability (QMG reduction of 0-2 points, grey), or improvement (QMG reduction ≥ 3 points; green). **(C)** shows mean MG-QoL15 trajectories (left) alongside the best individual improvement and proportion reaching a PASS (MG-QoL15 score ≤ 8; right). **(D)** reports steroid intake, displaying mean daily prednisone at BL and months 1, 3 and 6 (left) together with the maximal individual dose reduction within six months (right). In **(E)**, pyridostigmine dosing is depicted, showing mean daily doses at BL and at months 1,3, as well as 6 (left), together with the greatest individual dose reduction within six months (right). Error bars represent mean ± SD. Groups were compared with two-sided Student’s t-tests; p < 0.05 was considered significant. *BL, baseline; MG-ADL, Myasthenia Gravis Activities of Daily Living; QMG, Quantitative Myasthenia Gravis; PASS, Patient Acceptable Symptom State; SD, standard deviation*.

## Notes

### Competing Interest Statement

The authors have declared no competing interest.

### Funding Statement

This work was funded by the Else Kroener-Fresenius-Stiftung to CN (2023_EKEA.38). Further, this work was supported by the internal research funding program of the Heinrich-Heine University to MO and CN. Further study funding by the DGM (Deutsche Gesellschaft fuer Muskelkranke) to TR and to CN, by the German Research Foundation (DFG, to CN: NE 2774/2-1, to TR: 549557400/RU 2169/5-1, 417677437/GRK2578).

### Author Declarations

Etthics committee of Charite – Universitaetsmedizin Berlin gave ethical approval for this work (EA1/214/18).

