## Supplementary figures and images for "Early Versus Delayed Add-on Therapy in Generalized Myasthenia Gravis: A Multicenter Real-World Cohort Study"

### Supplemental Material 1

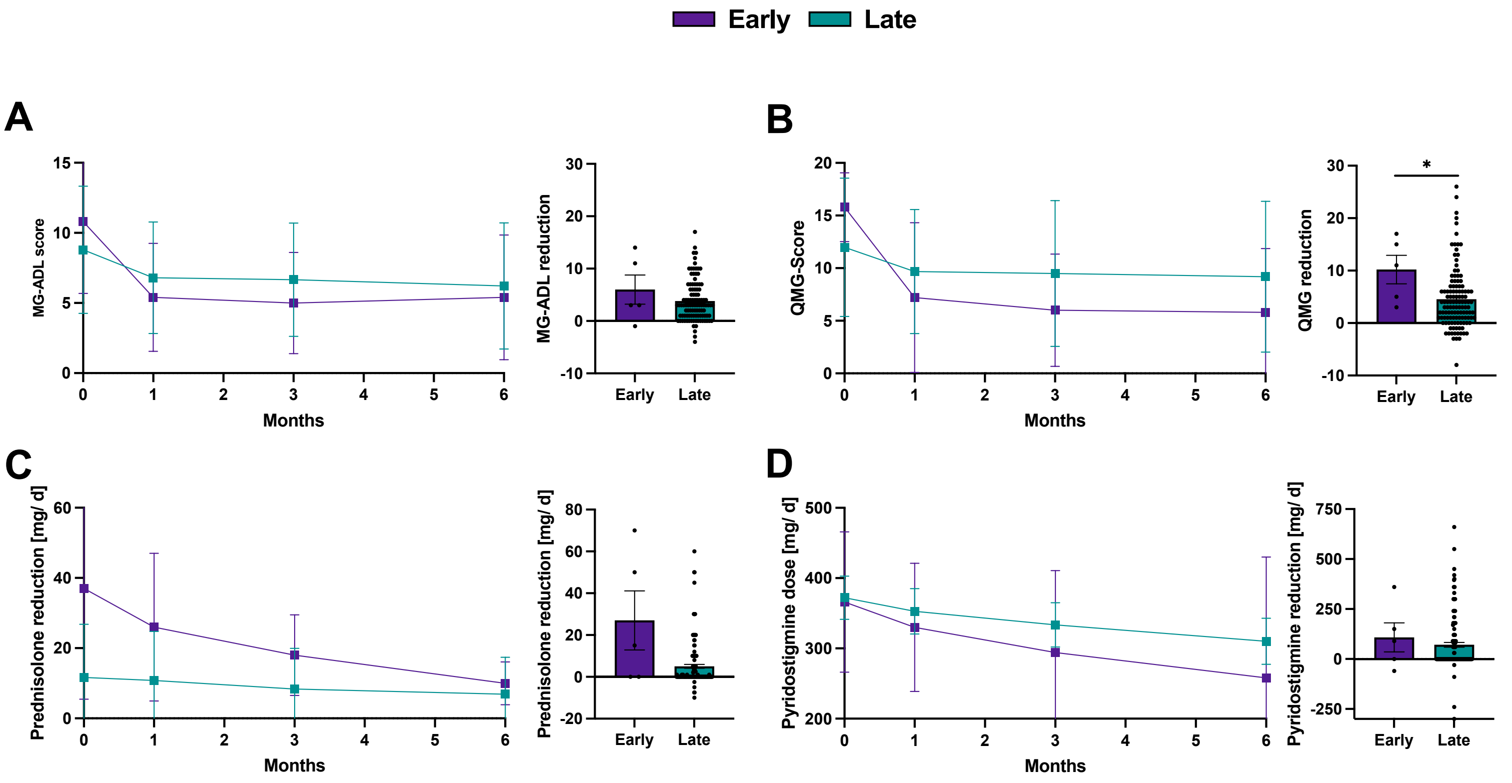


**Supplemental Figure 1**

### Supplemental Material 2

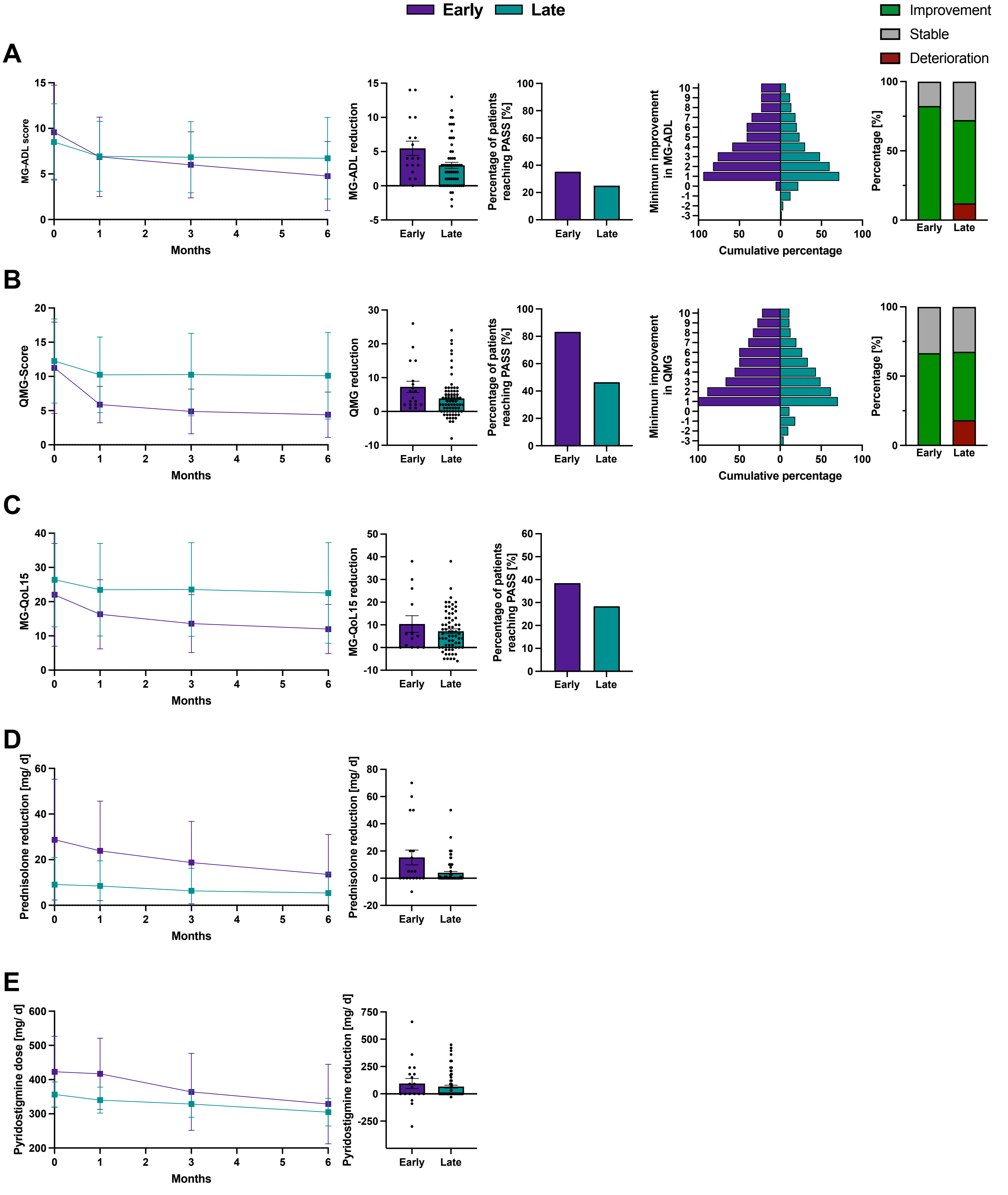


**Supplemental Figure 2**

### Supplemental Material 3

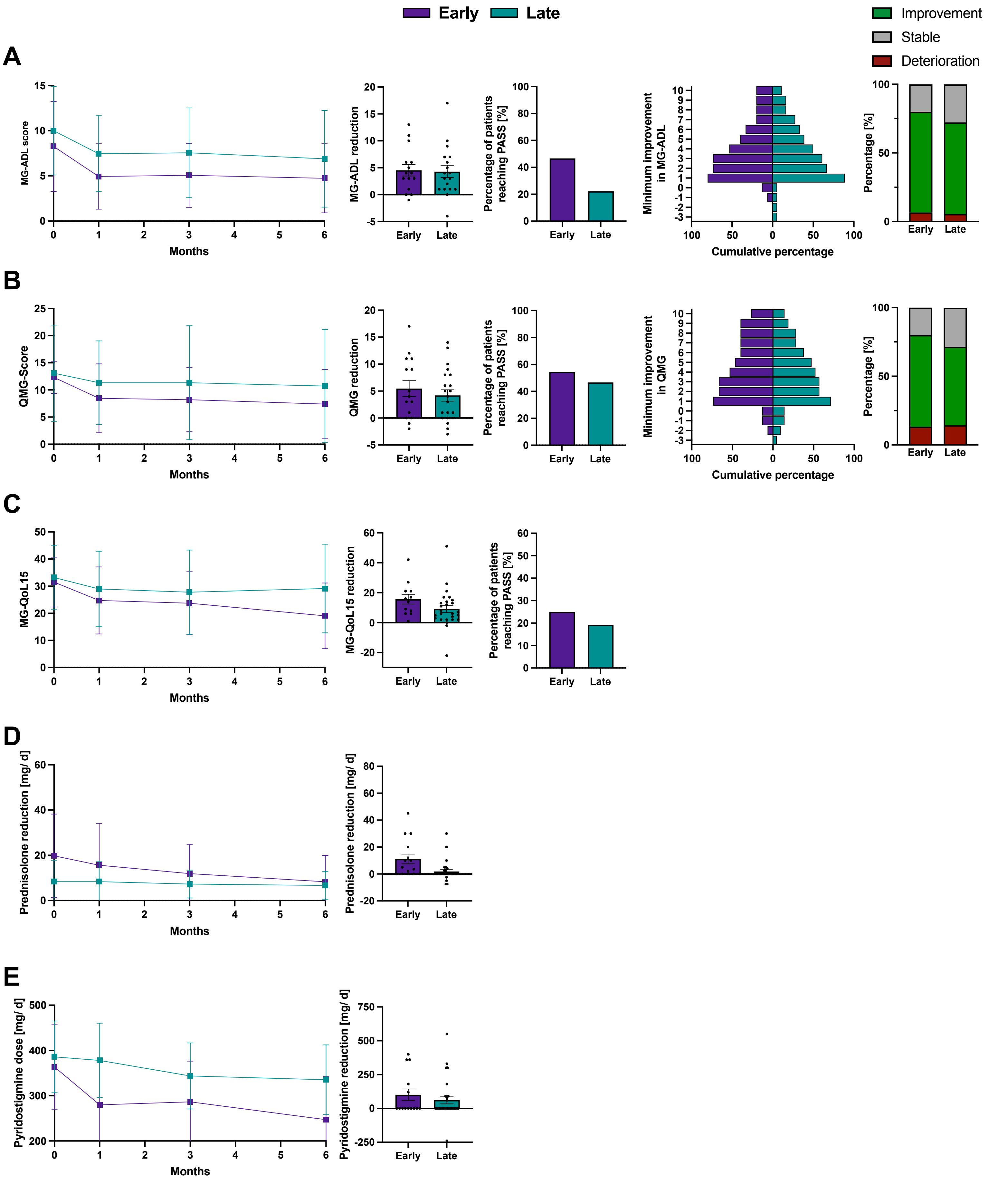


Supplemental Figure 3
